# Exome sequencing identifies rare coding variants in 10 genes which confer substantial risk for schizophrenia

**DOI:** 10.1101/2020.09.18.20192815

**Authors:** Tarjinder Singh, Benjamin M. Neale, Mark J. Daly, on behalf of the Schizophrenia Exome Meta-Analysis (SCHEMA) Consortium

**Author notes:** These authors contributed equally to this work. Correspondence should be addressed to: M.J.D., B.M.N., T.S.

## Abstract

By meta-analyzing the whole-exomes of 24,248 cases and 97,322 controls, we implicate ultra-rare coding variants (URVs) in ten genes as conferring substantial risk for schizophrenia (odds ratios 3 - 50, P < 2.14 × 10^-6^), and 32 genes at a FDR < 5%. These genes have the greatest expression in central nervous system neurons and have diverse molecular functions that include the formation, structure, and function of the synapse. The associations of NMDA receptor subunit *GRIN2A* and AMPA receptor subunit *GRIA3* provide support for the dysfunction of the glutamatergic system as a mechanistic hypothesis in the pathogenesis of schizophrenia. We find significant evidence for an overlap of rare variant risk between schizophrenia, autism spectrum disorders (ASD), and severe neurodevelopmental disorders (DD/ID), supporting a neurodevelopmental etiology for schizophrenia. We show that protein-truncating variants in *GRIN2A, TRIO*, and *CACNA1G* confer risk for schizophrenia whereas specific missense mutations in these genes confer risk for DD/ID. Nevertheless, few of the strongly associated schizophrenia genes appear to confer risk for DD/ID. We demonstrate that genes prioritized from common variant analyses of schizophrenia are enriched in rare variant risk, suggesting that common and rare genetic risk factors at least partially converge on the same underlying pathogenic biological processes. Even after excluding significantly associated genes, schizophrenia cases still carry a substantial excess of URVs, implying that more schizophrenia risk genes await discovery using this approach.

## Introduction

Schizophrenia is a severe psychiatric disorder with signs and symptoms that include hallucinations, delusions, disorganized speech and behavior, diminished emotional expression, social withdrawal, and cognitive impairment. The disorder has a lifetime risk of ~0.7%, is often disabling, and reduces life expectancy by nearly 15 years^1,2^. Existing therapies largely address primarily positive symptoms (e.g., hallucinations and delusions) and response to existing antipsychotic medications is highly variable with ~30% of patients classified as treatment resistant^3^. The lack of progress in therapeutic development is in part a consequence of our limited understanding of the molecular etiology of psychiatric disorders^3,4^.

It is well-established that schizophrenia has a substantial genetic component with contributions from across the allele frequency spectrum^5–8^. As initially theorized, the high heritability, consistency of prevalence across populations and increasing risk observed for individuals in more densely affected families suggested that polygenic predisposition should play a dominant role in defining schizophrenia risk in the population^1,9^. This has been borne out by genome-wide association studies (GWAS) which have now, in a companion paper, identified 270 common (minor allele frequency [MAF] > 1%) risk loci of individually small effect (median odds ratio [OR] < 1.05)^10^. As a class of variation, common variants explain ~24% of the variance in disease liability^11^. Several rare (MAF < 0.1%) recurrent copy number variants (CNVs) have also been robustly associated with schizophrenia, as exemplified by the dramatically higher rates of schizophrenia in 22q11.2 deletion carriers^7,12^. This suggests a role for rare gene-disrupting mutations with much larger effects on individual risk (OR 2 - 60). Because these are generally rare and often *de novo* mutations, they explain little of the genetic heritability, though they represent the strongest individual risk factors identified to date. Despite these successes in locus discovery, it remains challenging to move from individual associations to specific genes and disease mechanisms. Because causal variants in schizophrenia GWAS are predominantly non-coding, challenges related to fine-mapping and interpretation of intergenic and intronic elements limit our ability to confidently identify underlying genes, infer the mechanism by which they influence disease risk, and determine the direction of effect. CNVs of large effect, on the other hand, often disrupt hundreds of kilobases of the genome and multiple genes simultaneously, limiting our ability to derive clear functional insights^7^.

Analyzing rare coding variants offers a powerful complementary approach to identify genes in complex traits. Theory predicts that the forces of natural selection will tend to keep large effect risk variants at much lower frequencies in the population, especially in disorders such as schizophrenia that are associated with reduced fecundity^13^. However, most rare variants will have little or no functional consequence or impact on risk, posing a significant challenge in identifying those that are truly causal and complicating required analyses in which rare variants are tested as a group rather than individually. The most natural grouping for rare variants is within a gene, based on predicted functional consequence or evidence for deleteriousness^13,14^. As these variants are each seen in very few individuals - and often *de novo* or very recent - linkage disequilibrium can be readily avoided as a driver of spurious signals meaning that genes frequently can be identified directly. Furthermore, protein-truncating variants (PTVs) are among the most interpretable associations as they suggest that the effect on disease most commonly tracks with decreasing expression of the gene^15^. Earlier schizophrenia sequencing studies have established that ultra-rare and *de novo* mutations contribute to risk as a category, and have prioritized disease-relevant tissues and processes, specifically observing an enrichment in neuronal genes and synaptic processes^6,8,16-20^. Furthermore, these risk alleles are concentrated in genes with a near-complete depletion of protein-truncating variants in population studies, a result shared with other neurodevelopmental disorders^6,8^ and suggesting strong direct selection against such mutations. However, the analysis of URVs has had limited success in delivering individual gene discovery in schizophrenia because of power limitations, with only a single gene, *SETD1A*, identified as robustly associated^13,18^.

The Schizophrenia Exome Sequencing Meta-Analysis (SCHEMA) Consortium was formed as a global collaborative effort to analyze sequence data from many studies to advance gene discovery. Here, we generated, aggregated, harmonized variant identification, and meta-analyzed the exome sequences of 24,248 individuals with schizophrenia and 97,322 controls from seven continental populations. This analysis is, to our knowledge, one of the largest sequencing studies of a complex trait to date. As predicted by apparent rare variant burden in schizophrenia, increasing the sample size has led to the identification of 10 genes with URVs that confer substantial risk at exome-wide significance. Combining these findings with other large-scale sequencing studies, we find shared and distinct genetic signals between schizophrenia and other neurodevelopmental disorders. In tandem with a companion paper from the Psychiatric Genomics Consortium^10^, we provide evidence that common and ultra-rare coding variants identify an overlapping set of genes. Finally, we demonstrate that increased scale following this approach will uncover additional risk genes and help complete the genetic architecture of schizophrenia.

## Results

### Data description, generation, and quality control

We aggregated exome sequence data consisting of 24,248 individuals diagnosed with schizophrenia and 50,437 individuals without a known psychiatric diagnosis, recruited in eleven global collections that had previously contributed to common variant association efforts (Supplementary Methods, Figure 1A, Table S1). The sequence data for 7,979 cases had been previously presented in earlier publications^6,8,16–19^, while the remaining 16,269 cases are presented here for the first time. To ensure calibrated analyses, these samples were included in joint re-processing and variant calling using a standardized BWA-Picard-GATK pipeline as part of the larger Genome Aggregation Database (gnomAD) effort (Supplementary Methods); consequently, SCHEMA case-control samples with appropriate permissions are also included in the gnomAD v2 release^21^. After extracting SCHEMA samples from this callset, we performed quality control steps to ensure high quality of sequence data, exclude contaminated samples, identify parent-proband trios and other related individuals, and infer global ancestries (Supplementary Methods, Figure 1B, Figures S1-7, Table S2). We subsequently applied site- and genotype-level filters to generate a robust set of coding SNPs and indels for a well-matched case-control analysis (Supplementary Methods). Previous studies have shown that PTVs are concentrated in 3,063 genes under strong constraint in schizophrenia cases compared to controls^8,22^, and we replicated this result with consistent signals across our major cohorts (*P*_meta_ = 7.6 × 10^-35^; OR = 1.26, 95% CI = 1.22 - 1.31, Figure 1C, Figure S8).

**Figure 1.**
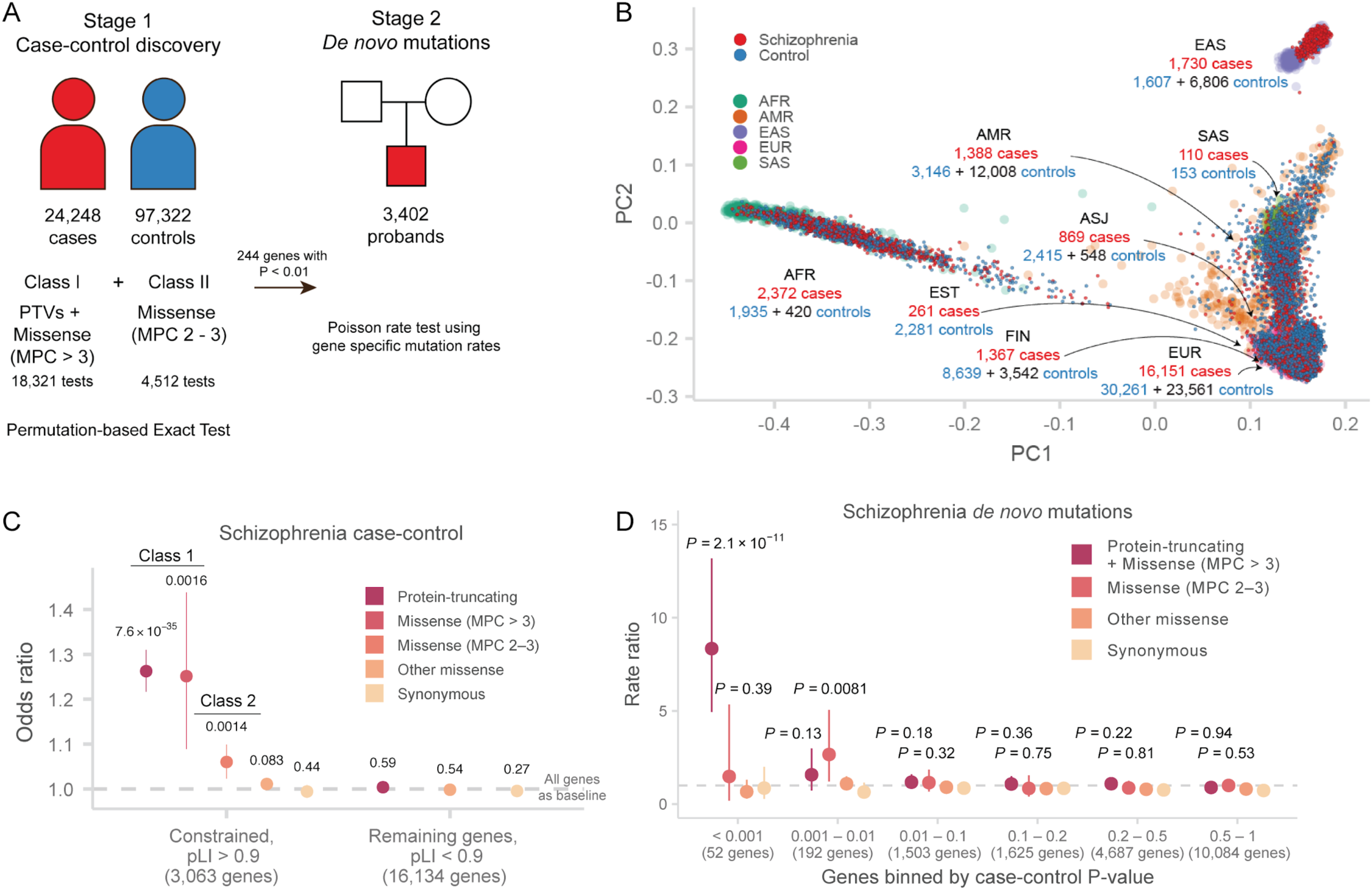
Study design and analytic approach. **A:** Study design. Case-control and parent-proband trio sample sizes, variant classes, and analytical methods are described. The case-control stage is shown on the left, and the *de novo* mutation stage is shown on the right. **B**: Principal components analysis of SCHEMA samples. 1000 Genomes samples with reported ancestry are plotted in the background, and SCHEMA samples are displayed in the foreground. For each global ancestry group, we report the number of cases and controls in the discovery data set in red and blue respectively, and the number of external controls in black. AFR: African, ASJ: Ashkenazi Jewish, AMR: Latin American, EAS: East Asian, EST: Estonian, FIN: Finnish, EUR: non-Finnish European, SAS: South Asian. **C:** Case-control enrichment of ultra-rare protein-coding variants in genes intolerant of protein-truncating variants. *P* values displayed are from comparing the burden of variants of the labeled consequence in cases compared to controls. By definition, MPC enrichment is only shown for pLI > 0.9 genes. Bars represent the 95% CIs of the point estimates. pLI: probability of loss-of-function intolerant in the gnomAD database. **D:** Enrichment of schizophrenia *de novo* mutations in *P* value bins derived from the Stage 1 (case-control) gene burden analysis. The enrichment *P* values displayed are calculated as a Poisson probability having equal or greater than the observed number of mutations given the baseline mutation rate in 3,402 schizophrenia trios. The relative rate is given by the ratio of observed to expected rate of *de novo* mutations. Bars represent the 95% CIs of the point estimates.

### Analysis approach

To increase power for gene discovery, we incorporated variant counts from additional samples from non-psychiatric and non-neurological collections that were aggregated as part of the gnomAD consortium effort (Supplementary Methods)^21^. We attempted to control for technical and methodological batch effects that may arise from this approach in both variant calling and additionally via permutation testing described below. All samples in gnomAD and SCHEMA consortia were re-processed and joint called using the same pipeline, and the same variant filters were applied to arrive high-quality calls. Importantly, we restricted our analysis to coding exons with high-quality data across all major exome capture technologies, reducing any artifacts that may arise from coverage differences(Supplementary Methods, Figures S1-2). After incorporating variant counts from additional 46,885 gnomAD controls, our combined discovery data set is composed of 24,248 cases and 97,322 population controls (Figure 1A, 1B, Table S3).

Because only summary-level variant counts were available for the 46,885 external controls, we tested for an excess of disruptive variants per gene using a Fisher’s exact test in which statistical significance was determined by case-control permutations within each strata (Supplementary Materials, Table S3). As in other sequencing studies, we enriched for pathogenic variants by restricting our analysis to ultra-rare variants (defined as minor allele count [MAC] ≤ 5) that are also either PTVs (defined as stop-gained, frameshift, and essential splice donor or acceptor variants) or damaging missense variants as defined by the MPC pathogenicity score^23,24^(Supplementary Methods). We found that missense variants with MPC > 3 have a global signal on par with PTVs in schizophrenia, autism spectrum disorders, and severe neurodevelopmental disorders, while variants with MPC 2 - 3 has a significant but weaker signal than PTVs and were therefore analyzed separately (Figure 1C, Figures S9, S10, S11, Table S4, Supplementary Methods). Motivated by these observations, we performed a burden test of PTVs and MPC > 3 variants (Class I) to generate a *P* value for 18,321 protein-coding genes (Supplementary Methods). In the 4,512 genes with MPC 2 - 3 (Class II) variants, we perform an additional test aggregating these variants, and meta-analyze these gene statistics with Class I *P* values using a weighted Z-score method (Supplementary Methods). To ensure the robustness of the results generated by this approach, we observed the expected null distribution of *P* values in gene-based tests of synonymous variants in each strata and in the meta-analysis (Figure S12, S13). Additionally, we observed no inflation of synonymous *P* values using the Mantel-Haenszel test even after limiting our analysis to genes with larger total numbers of alleles (gene-wide MAC > 10, 50, or 100), where we had greater power to detect potential artifacts (Figure S14, S15).

Previous studies had integrated case-control and trio-based *de novo* mutations for gene discovery^18,24^, and to this end, we aggregated and re-annotated *de novo* mutations from 3,402 published parent-proband trios (Supplementary Methods). Despite the sizable number of trios, there were few *de novo* mutations for analysis with only 325 genes with one or more *de novo* PTV and only 449 with at least one Class I or Class II mutations. Using Poisson rate tests based on expected mutation rate^25^, we found these *de novo* mutations are enriched for the 244 genes with *P* < 0.01 in our case-control analysis (Figure S16, Table S5), with limited or no signal in the remaining genes in the genome (Figure 1D). The most striking enrichment was observed for the 52 genes with case-control *P* < 0.001 (Class I mutations: *P* = 2.1 × 10^-11^; Rate ratio = 8.3, 95% CI = 4.9 - 13), which provides additional reassurance of the robustness of our case-control gene results. Motivated by these observations, we calculated *de novo* Class I and II *P* values in the 244 genes with *P*_case-control_ < 0.01 using the Poisson rate test and meta-analyzed them with our case-control test statistic using a weighted Z-score method to increase power (Supplementary Methods).

### Risk genes implicated by rare protein-coding variants

Combined, our meta-analysis of 24,248 cases, 97,322 controls, and *de novo* mutations from 3,402 trios implicates 10 genes in which ultra-rare coding variants are significantly associated with schizophrenia (P < 2.14 × 10^-6^ corresponding to 0.05/23,321 tests; Figure 2A, 2B). These top associations as a group are supported by complementary types of variation that include case-control PTVs, damaging missense variants, and *de novo* mutations (Table 1, Table S5). Although confidence intervals are wide, URVs in these genes appear to confer substantial risk, with odds ratio of PTVs and Class I variants ranging from 3 to 50. As expected, all ten genes are among the most constrained genes in the genome, with a substantial depletion of PTVs compared to chance expectation^21^. The annotated functions of these genes are diverse and include ion transport *(CACNA1G, GRIN2A*, and *GRIA3)*, neuronal migration and growth *(TRIO)*, transcriptional regulation *(SP4, RB1CC1*, and *SETD1A)*, nuclear transport *(XPO7)*, and ubiquitin ligation *(CUL1, HERC1)*. We include a brief discussion of the known biological functions of these genes in Box 1. Beyond these ten genes, we identify 22 additional genes at a False Discovery Rate (FDR) < 5% (Figure 2A, Table S5). We observe notable deviation at the tail of the distribution beyond the associated genes, suggesting that more genes remain to be discovered (Figure 2B). We report all high-quality variants, relevant annotations, and gene-level results on a public browser at https://schema.broadinstitute.org.

**Figure 2.**
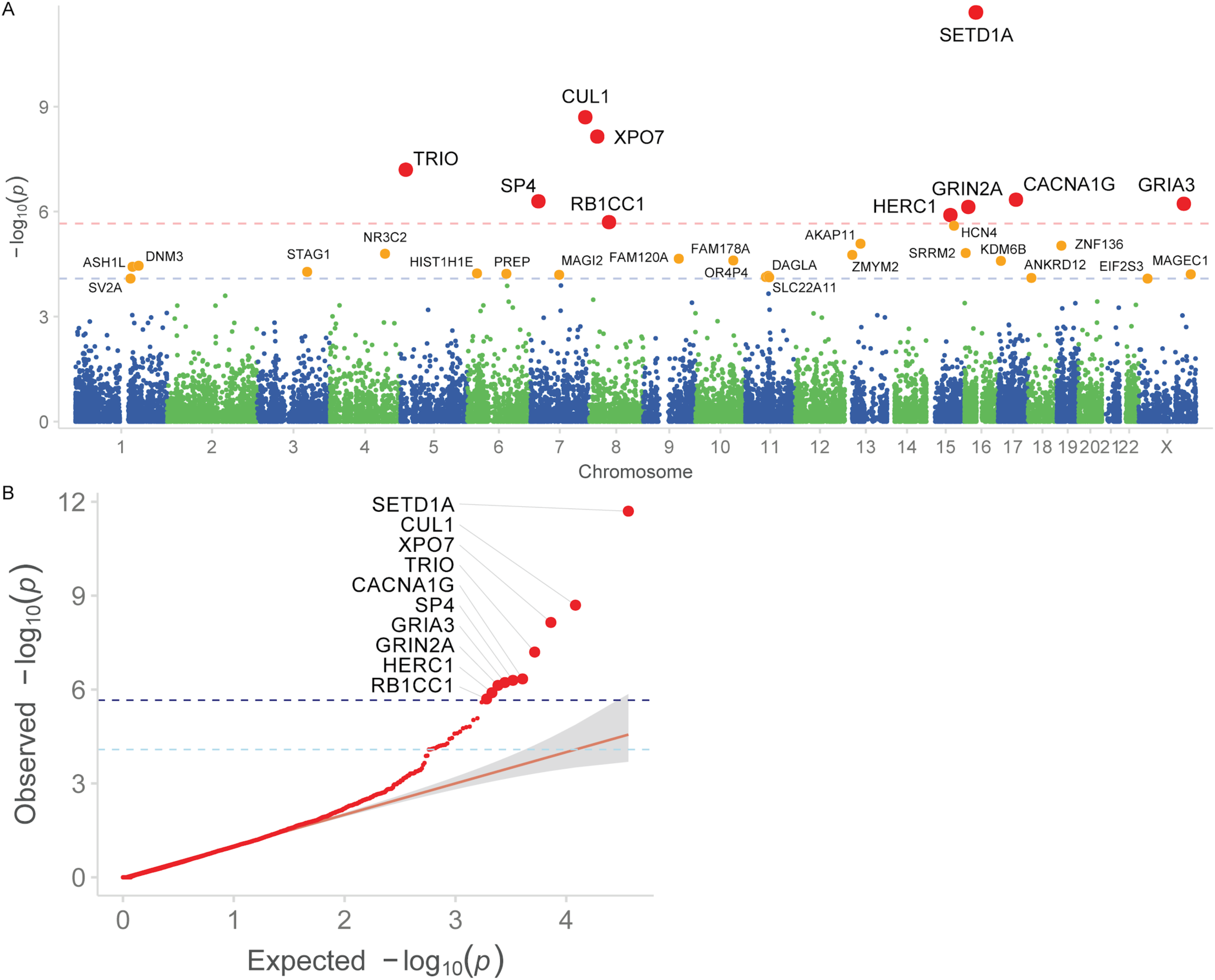
Results from the meta-analysis of ultra-rare coding variants in 3,402 trios, 24,248 cases, and 97,322 controls. **A:** Manhattan plot. –log_10_ *P*-values are plotted against the chromosomal location of each gene. Genes reaching exome-wide significance are in red, and genes significant at FDR < 5% are in orange. Red dashed line: *P* = 2.14 × 10^-6^; Blue dashed line: FDR < 5%, or *P =* 8.23 × 10^-5^. **B:** Q-Q plot. Observed –log_10_ *P*-values are plotted against expectation given a uniform distribution. Genes reaching exome-wide significance are plotted with a larger size. The direction of effect is indicated by the color of each point. Dark blue dashed line: *P* = 2.14 × 10^-6^; Light blue dashed line: FDR < 5%.

**Table 1.**
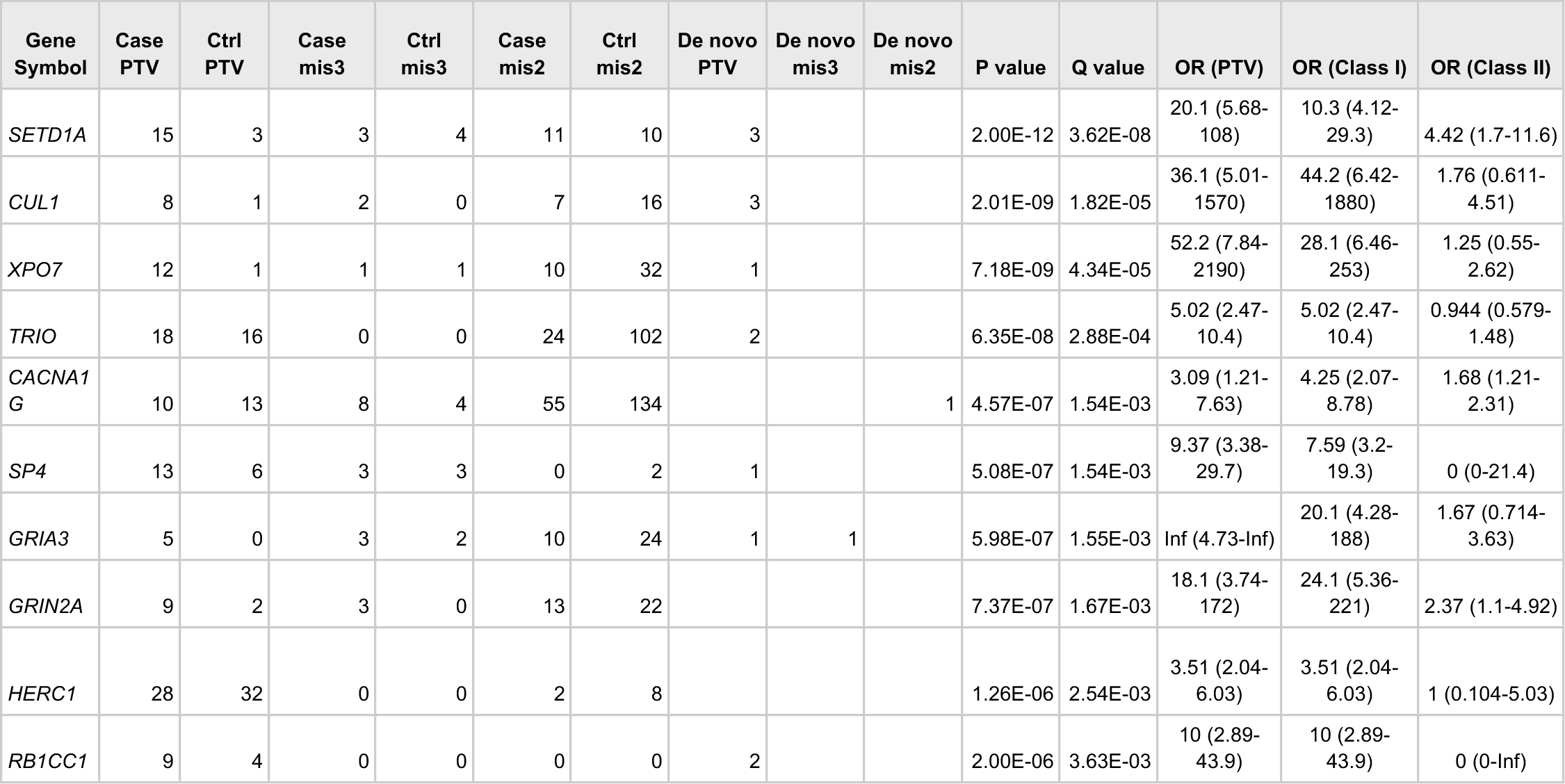
Case-control and *de novo* counts of the ten Bonferroni significant genes in the main analysis. Case-control counts displayed are the total counts for variants with minor allele count <= 5. PTV: protein-truncating variant, mis3: missense variants with MPC > 3, mis2: missense variants with MPC 2 - 3; Q value: adjusted *P* value after FDR adjustment; Class I: PTV and missense variants (MPC > 3); Class II: missense variants (MPC 2 - 3).

| Gene Symbol | Case PTV | Ctrl PTV | Case mis3 | Ctrl mis3 | Case mis2 | Ctrl mis2 | De novo PTV | De novo mis3 | De novo mis2 | P value | Q value | OR (PTV) | OR (Class I) | OR (Class II) |
| --- | --- | --- | --- | --- | --- | --- | --- | --- | --- | --- | --- | --- | --- | --- |
| SETD1A | 15 | 3 | 3 | 4 | 11 | 10 | 3 |  |  | 2.00E-12 | 3.62E-08 | 20.1 (5.68-108) | 10.3 (4.12-29.3) | 4.42 (1.7-11.6) |
| CUL1 | 8 | 1 | 2 | 0 | 7 | 16 | 3 |  |  | 2.01E-09 | 1.82E-05 | 36.1 (5.01-1570) | 44.2 (6.42-1880) | 1.76 (0.611-4.51) |
| XPO7 | 12 | 1 | 1 | 1 | 10 | 32 | 1 |  |  | 7.18E-09 | 4.34E-05 | 52.2 (7.84-2190) | 28.1 (6.46-253) | 1.25 (0.55-2.62) |
| TRIO | 18 | 16 | 0 | 0 | 24 | 102 | 2 |  |  | 6.35E-08 | 2.88E-04 | 5.02 (2.47-10.4) | 5.02 (2.47-10.4) | 0.944 (0.579-1.48) |
| CACNA1G | 10 | 13 | 8 | 4 | 55 | 134 |  |  | 1 | 4.57E-07 | 1.54E-03 | 3.09 (1.21-7.63) | 4.25 (2.07-8.78) | 1.68 (1.21-2.31) |
| SP4 | 13 | 6 | 3 | 3 | 0 | 2 | 1 |  |  | 5.08E-07 | 1.54E-03 | 9.37 (3.38-29.7) | 7.59 (3.2-19.3) | 0 (0-21.4) |
| GRIA3 | 5 | 0 | 3 | 2 | 10 | 24 | 1 | 1 |  | 5.98E-07 | 1.55E-03 | Inf (4.73-Inf) | 20.1 (4.28-188) | 1.67 (0.714-3.63) |
| GRIN2A | 9 | 2 | 3 | 0 | 13 | 22 |  |  |  | 7.37E-07 | 1.67E-03 | 18.1 (3.74-172) | 24.1 (5.36-221) | 2.37 (1.1-4.92) |
| HERC1 | 28 | 32 | 0 | 0 | 2 | 8 |  |  |  | 1.26E-06 | 2.54E-03 | 3.51 (2.04-6.03) | 3.51 (2.04-6.03) | 1 (0.104-5.03) |
| RB1CC1 | 9 | 4 | 0 | 0 | 0 | 0 | 2 |  |  | 2.00E-06 | 3.63E-03 | 10 (2.89-43.9) | 10 (2.89-43.9) | 0 (0-Inf) |

The identification of individual genes provides support for more specific mechanistic hypotheses underlying schizophrenia pathogenesis. Developed from neuropharmacological and neuropathological observations, the glutamatergic hypothesis postulates that the hypofunction of glutamatergic signaling through NMDA receptors is a possible mechanism of disease^26^ (Box 1). Here, we find that PTV and damaging missense variants in NMDA receptor subunit *GRIN2A* confer substantial risk for schizophrenia (*P* = 7.37 × 10^-7^; Class I [PTV and MPC > 3] OR 24.1, 95% CI 5.36 - 221; Class II [MPC > 2] OR 2.37, 95% CI 1.1 - 4.92). Schizophrenia GWAS also identified a common variant at *GRIN2A* (OR = 1.057, *P =* 1.57 × 10^10^), providing an allelic series in which different perturbation of gene function results in severity of disease risk (Figure 3A)^5^.The NMDA receptor changes in composition during prenatal to postnatal neurodevelopment with *GRIN2A* predominantly expressed during late childhood and adolescence, recapitulating expected epidemiological observations on schizophrenia age-of-onset (Supplementary Information, Figure 3B)^27^. We additionally find that risk URVs in AMPA receptor subunit *GRIA3* confer substantial risk (*P* = 5.98 × 10^-7^; Class I [PTV and MPC > 3] OR 20.1 95% CI 4.28 - 188; Table 1). Combined, our results from exome sequencing support the dysregulation of the glutamatergic system as a mechanistic hypothesis for the development of schizophrenia, and that the specific identification of genes by coding variation may provide new avenues of understanding disease pathogenesis.

**Figure 3.**
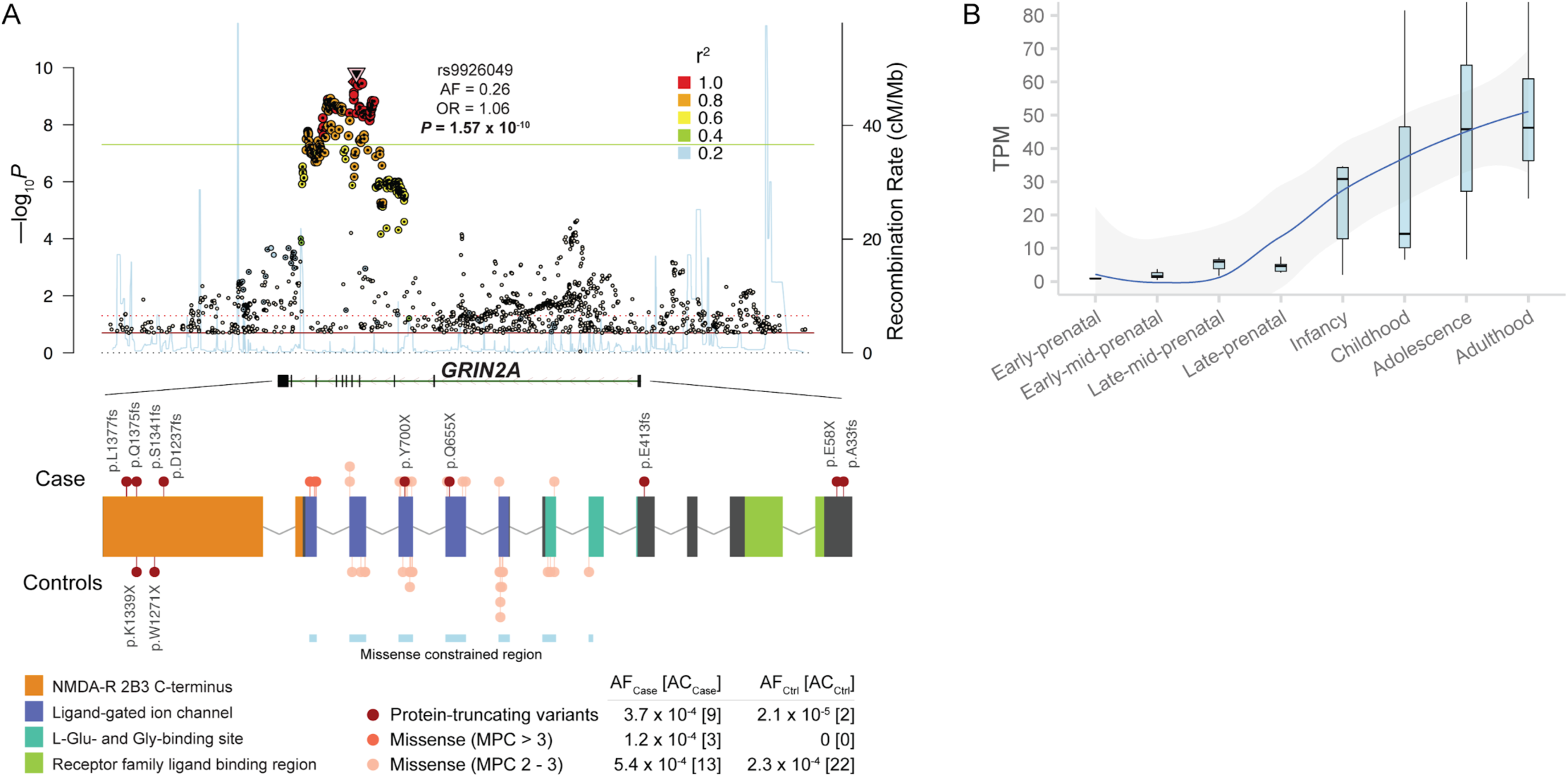
Biological insights from exome sequence data. **A:** Common and rare allelic series at NMDA receptor subunit *GRIN2A*. The Locus Zoom plot (top) displays the common variant association of the gene. The color of each dot corresponds to the LD with the index SNP, and the properties of the index SNP are displayed. The gene plot (bottom) displays the protein-coding variants that contribute to the exome signal in *GRIN2A*. Variants discovered in cases are plotted above the gene, and those from control are plotted below. Each variant is colored based on inferred consequence, and the protein domains and missense constrained regions of the gene are also labelled^23,28^. The frequencies and counts in cases and controls are displayed for each variant class. AF: allele frequency, AC: allele count. **B:** Temporal expression of *GRIN2A* in the human brain. We show *GRIN2A* expression in four prenatal and four postnatal periods derived from whole-brain tissue in BrainSpan^27^. The expression values plotted are in transcript-per-million (TPM).

### Shared genes and processes identified by common variants and ultra-rare protein-coding variants

Pathway analyses of common variants have prioritized disease-relevant tissues and cell types, and in some cases, independently recapitulating known biology^5,29,30^. To derive insights from global patterns of rare coding variants, we tested for an excess burden of URVs in schizophrenia cases compared to controls in 1,732 broadly-defined gene sets from databases of biological pathways (e.g. Gene Ontology, REACTOME, KEGG) and experimental data (Supplementary Information)^8^. We observed significant enrichment of URVs in 33 gene sets *(P* < 2.9 × 10^-5^) that recapitulated consistent and overlapping cellular compartments and biological processes, including definitions of the postsynaptic density (human cortex biopsy post-synaptic density; *P* = 1.2 × 10^-12^), chromatin modification (GO:0016568; *P* = 1.8 × 10^-12^), regulation of ion transmembrane transport (GO:0034765; P = 6.7× 10^-7^), axon guidance (*P* = 5.4 × 10^-6^), voltage-gated cation channel activity (GO:0022843; P = 8.1 × 10^-6^), and synaptic transmission (GO:0007268; *P* = 1.79 × 10^-5^) (Table S6, Figure S18). Because of the clear synaptic signal, we investigated in the refined synaptic ontology defined by the SynGO consortium^31^, and found consistent enrichment for postsynaptic components and processes (GO:0098794; *P* = 3.9 × 10^6^; Table S7). These global observations are consistent with the known functions of the individual risk genes now implicated by rare variation (Box 1). Following earlier reports studying heritability enrichment in GTEx tissues^5,30^, we found that genes with the highest specific expression in brain regions showed the strongest enrichment of risk URVs, most significantly in the human frontal cortex (P = 1.63 × 10^-8^) and with limited signal in the other tissue types (Figure S19, Table S8, S9). To further deconvolute this signal, we investigated which single cell types in the mouse nervous system show the highest specific expression for the 32 (FDR <5%) schizophrenia risk genes (Supplementary Information)^32,33^. Here, we found widespread enrichments across central nervous system neurons with limited to no signal in glial cells and peripheral nervous system neurons (Table S10, Figure S20). Thus, at a high level, global analysis of ultra-rare protein-coding variation independently recapitulated known biology related to schizophrenia pathogenesis, including processes, cellular components, and tissues previously implicated by common variant analyses.

To evaluate the overlap of schizophrenia associations from common variants and ultrarare coding variant analyses, we jointly analyzed our results with the largest GWAS of schizophrenia to date, which identified common variant associations at 270 distinct loci from the analysis of 69,369 cases and 236,642 controls^10^. Statistical fine-mapping prioritized the likely underlying protein-coding gene at 69 of these associations (Table S11), and we found a case-control enrichment of URVs in these genes (*P*_meta_ = 1.1 × 10^-3^; OR_Class I_ = 1.42, 1.17 - 1.73 95% CI; Figure 4A, Table S12). Beyond the statistical enrichment, *GRIN2A* and *SP4*, two of the ten significant rare variant genes, had clear associations in schizophrenia GWAS (Figure 3A, Figure 4B). Furthermore, *FAM120A* and *STAG1* resided in more complex GWAS-associated regions containing multiple genes but were prioritized among their neighbors as FDR < 5% in our sequencing study (Figure 4C, 4D). Combined, these results suggest there is at least partial convergence in the genes and biological processes implicated by common and ultra-rare genetic variation, and that ultra-rare coding variants can be leveraged to prioritize genes within GWAS loci.

**Figure 4.**
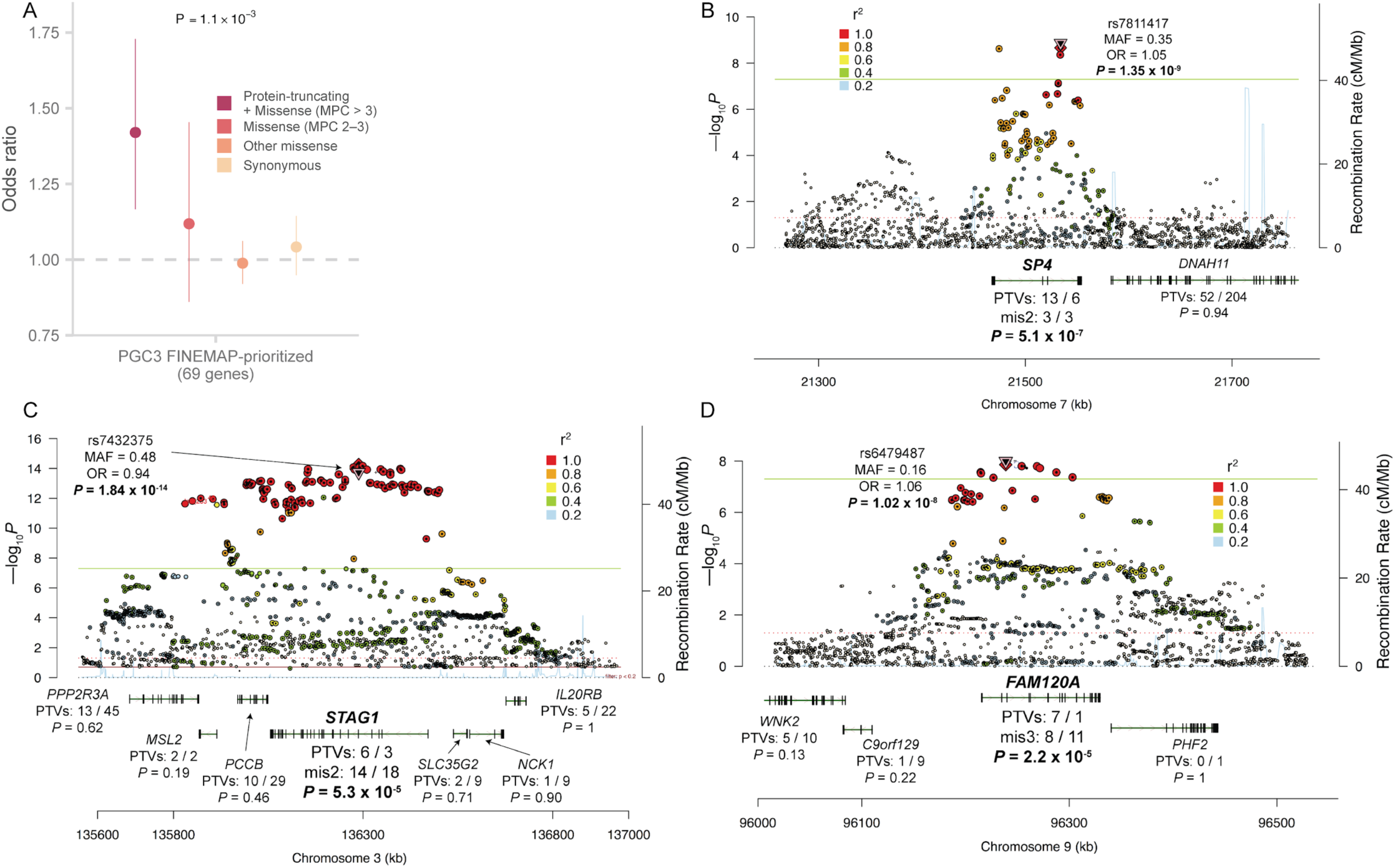
Shared genetic signal with schizophrenia GWAS. **A:** Case-control enrichment of ultra-rare protein-coding variants in genes prioritized from fine-mapping of the PGC schizophrenia GWAS^10^. The reported *P* value is the Fisher combined *P* value of Class I and Class II variants. Bars represent the 95% CIs of the point estimates. **B, C, D:** Prioritization of GWAS loci using exome data. The Locus Zoom plot of three GWAS loci is displayed. For each gene in or adjacent to the region, we show the case-control counts of PTVs in the exome data, along with the meta-analysis P-value. *SP4, STAG1* and *FAM120A* are highlighted as the only genes with notable signals in the exome data within each locus.

### Shared and distinct genetic risk with other neurodevelopmental disorders

Exome sequencing studies of autism spectrum disorders (ASD) and severe neurodevelopmental disorders (DD/ID) have leveraged ultra-rare coding variants to identify risk genes. These studies have established that the genetic signals were concentrated in constrained genes and shared between the two disorders^34,35^. Most recently, the analysis of *de novo* mutations from 31,058 DD/ID trios implicated 299 genes, while the analysis of 11,986 ASD cases identified 102 genes at FDR < 10% (Table S1 1)^24,36^. We found a significant excess of URVs in schizophrenia cases compared to controls in the 299 DD/ID-associated genes (*P*_meta_ = 1.5 × 10^-14^; OR_Class I_ = 1.44, 1.3 - 1.6 95% CI), and in the 102 ASD-associated genes (*P*_meta_ = 3.7 × 10^-7^; OR_Class I_ = 1.45, 1.23 - 1.72 95% CI; Figure 5A; Table S12). Thus, some schizophrenia rare variant risk appears to be shared with other neurodevelopmental disorders.

**Figure 5.**
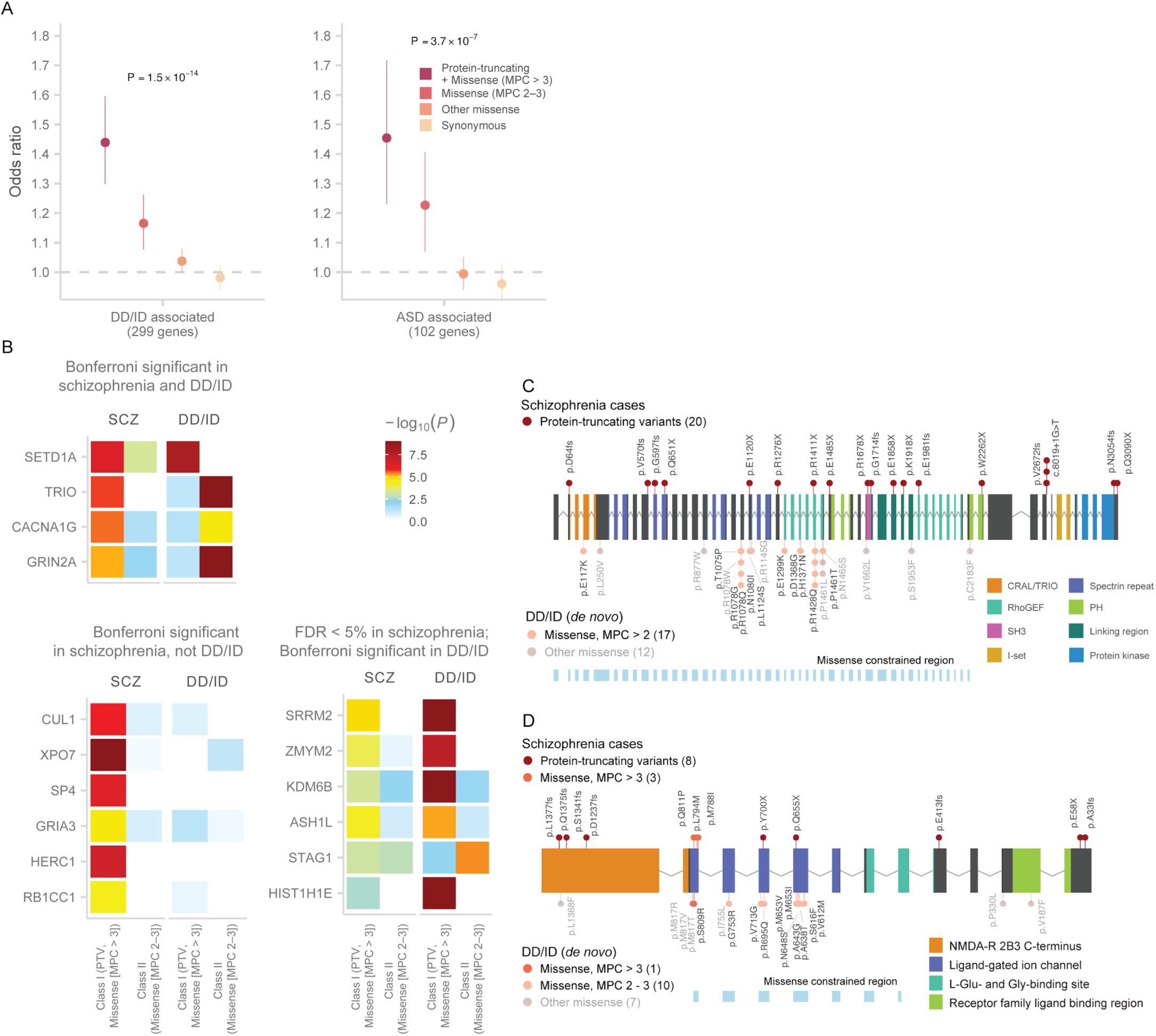
Shared genetic signal between schizophrenia and other neurodevelopmental disorders. **A:** Case-control enrichment of ultra-rare protein-coding variants in DD/ID and ASD-associated genes. We test for the burden of schizophrenia URVs in genes identified in the most recent exome sequencing studies of ASD and DD/ID^24 36^. The reported *P* value is the Fisher combined *P* value of Class I and Class II variants. Bars represent the 95% CIs of the point estimates. **B:** Heatmap displaying the strength of association for schizophrenia-associated genes in our discovery data set and in genes implicated by *de novo* mutations in trios diagnosed with DD/ID. We display three groups of genes: Bonferroni significant in schizophrenia and DD/ID, Bonferroni significant only in schizophrenia, and FDR < 5% in schizophrenia and Bonferroni significant in DD/ID. The degree of association from each sequencing study is displayed as the color corresponding to –log_10_ *P*-values in that study. The case-control P-value is reported for schizophrenia, while *de novo* enrichment from a Poisson rate test is reported for DD/ID. Results are further stratified to tests of Class I (PTV and MPC > 3) and Class II (missense [MPC 2 - 3]) variants. **C:** Allelic series in *TRIO* between schizophrenia and DD/ID risk variants. The gene plot displays the protein-coding variants that contribute to the exome signal in *TRIO*. Variants discovered in schizophrenia cases are plotted above the gene, and missense *de novo* mutations from DD/ID probands are plotted below. Each variant is colored based on inferred consequence, and the protein domains of the gene are also labelled. The variant counts are displayed for each variant class. **D:** Allelic series in *GRIN2A*. See **D** for description.

With 31,058 trios, the scale of gene discovery in severe DD/ID provided sufficient power to evaluate the individual schizophrenia risk genes associated in our study for a role in broader neurodevelopmental disorders. Nine of the ten schizophrenia genes showed limited *de novo* PTV signal in DD/ID, with a combined 8 *de novo* PTVs observed in these genes (X_exp_ = 4.98; *P*_pois_ = 0.13; Figure 5B; Table S13). *SETD1A* had a significant *de novo* PTV signal in DD/ID (X_obs_ = 8, X_exp_ = 0.41; *P* = 1.3 × 10^-8^), supporting an earlier report that described *SETD1A* as a gene associated with both schizophrenia and broader neurodevelopmental disorders^18^. We also observed a missense signal in *SETD1A* in our study (Table 1; Figure S21). Extending this analysis to the additional 22 FDR < 5% genes, we found that six genes *(STAG1, ASH1L, ZMYM2, KDM6B, SRRM2*, and *HIST1H1E)* were significantly associated with DD/ID in addition to schizophrenia (Figure 5B; Table S13). Among these FDR < 5% genes, *ASH1L, KDM6B* and *NR3C2* were associated with ASD ^24^ (Table S13). Broadly speaking, while PTV mutations in certain genes are joint risk factors for schizophrenia and DD/ID, the majority of schizophrenia associations reported here appear to have little or no role in DD/ID despite the enormous power of published DD/ID studies to date.

Notably, three of the ten risk genes for schizophrenia *(TRIO, GRIN2A*, and *CACNA1G)* were associated with risk of severe DD/IDs exclusively through *de novo* missense mutations that cluster within each gene (Figure 5B; Table S13), while the schizophrenia signal was largely driven by PTVs. *De novo* missense mutations in *TRIO* significantly disrupted the exons preceding or containing the RhoGEF domain (Figure 5C)^36,37^, and *de novo* missense mutations in *GRIN2A* cluster at the base of the ion channel with the most mutations in the exon encoding for the pore of the complex (Figure 5D). *STAG1*, which had a common and rare variant signal in schizophrenia (Figure 4D), was associated with DD/ID primarily through *de novo* missense mutations (Figure 5B; Table S13). These observations suggest schizophrenia and childhood onset neurodevelopmental disorders share some genes and biological processes, but that at least in some cases, the severity or the nature of the functional impairment differs between disorders.

We explored what properties may differ between schizophrenia- and DD/ID-associated risk genes, and hypothesized that DD/ID genes were under stronger evolutionary constraint with a bias towards prenatal expression when compared to schizophrenia genes. While schizophrenia genes (FDR < 5%) were under substantial genic constraint compared to expectation (Figure S22; M-W U test; *P* = 2.9 × 10^-7^; Supplementary Information), they are significantly less constrained than DD/ID-associated genes (M-W U test; *P* = 3.5 × 10^-5^). Furthermore, schizophrenia genes as a group did not show pre- or postnatal bias in brain expression (*P* = 0.21; Figure S23), while DD/ID-associated genes were overwhelmingly prenatal in expression (*P* = 7.5 × 10^-20^). Indeed, individual genes like *SETD1A, TRIO*, and *SP4* exhibited prenatal expression while *GRIN2A* and *GRIA3* showed postnatal expression (Figure S24). These observations offer the possibility that certain properties may differentiate genes for adult psychiatric disorders and more severe DD/IDs.

### Contribution of ultra-rare PTVs to schizophrenia risk

Efforts in the past decade are beginning to generate a more comprehensive view of the genetic architecture for schizophrenia, composed of common variants of small effects, large CNVs with elevated frequencies driven by genomic instability, and now, URVs of large effect implicating individual genes (Figure 6A)^5,7^. Because schizophrenia as a trait is under strong selection^38–40^, we expect that URVs of large effect to be frequently *de novo* or of very recent origin and contribute to risk in only a fraction of diagnosed patients. We quantified the contribution of PTVs to risk first in our full schizophrenia data set, and then partitioned the *de novo* and inherited contributions in 2,304 parent-proband trios. We restrict these analyses to the 3,063 PTV-intolerant (pLI > 0.9) genes in which schizophrenia risk URVs are concentrated. We observed 0.057 (0.049 - 0.065 95% CI) extra singleton PTV variants per individual in cases compared to controls, suggesting ~5.7% of cases carried a PTV relevant to disease risk. In the 2,304 trios, 0.0394 (0.014 - 0.065 95% CI; 74%) extra singleton PTV variants were inherited per proband, and 0.0121 (0.0022 - 0.02 95% CI; or 26%) extra *de novo* PTV mutations in constrained genes were identified in cases compared to controls. In contrast, DD/ID probands have 0.111 (0.103 - 0.119 95% CI) extra *de novo* PTV mutations in constrained genes, while ASD individuals have 0.0478 (0.0387 - 0.0568 95% CI) extra *de novo PTV* mutations (Figure S25; Supplementary Information). In the ten schizophrenia-associated genes, 7 *de novo* mutations and 13 transmitted variants are observed in 2,304 trios, suggesting that 0.86% of patients are carriers and ~35% of variants are *de novo*. Finally, the genome-wide signal in constrained genes (pLI > 0.9: OR = 1.26, *P* = 7.6 × 10^-35^) remains significant even after excluding the 32 FDR < 5% genes (OR = 1.23, *P* = 4.3 × 10^-27^; Figure 6B, Table S4), reaffirming the genetic heterogeneity underlying schizophrenia risk and suggesting that the majority of schizophrenia risk genes in which rare variants confer risk remain to be discovered.

**Figure 6.**
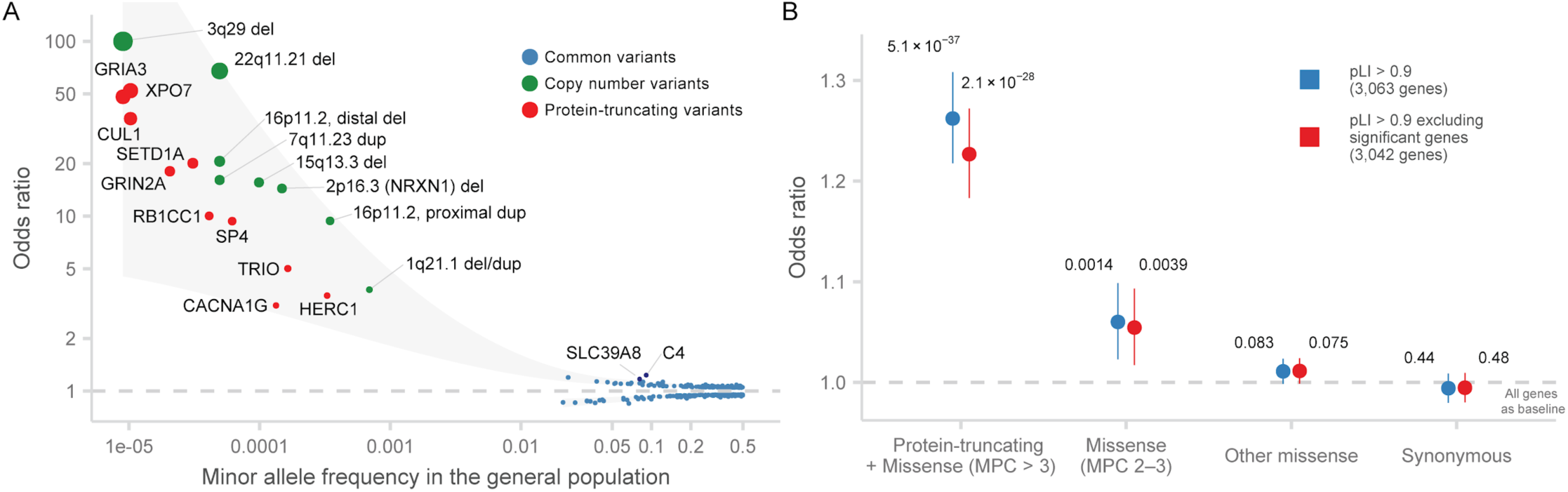
The contributions of ultra-rare PTVs to schizophrenia risk. **A:** Genetic architecture of schizophrenia. Significant genetic associations for schizophrenia from the most recent GWAS, CNV, and sequencing studies are displayed. The in-sample odds ratio is plotted against the minor allele frequency in the general population. The color of each dot corresponds to the source of the association, and the size of the dot to the odds ratio. The shaded area represented the loess-smoothed lines of the upper and lower bounds of the point estimates. **B:** Case-control enrichment of ultra-rare protein-coding variants in genes intolerant of protein-truncating variants after excluding schizophrenia-associate genes. We perform the test with all constrained genes (pLI > 0.9) and after excluding all schizophrenia-associated genes with FDR < 5%. *P* values displayed are from comparing the burden of variants of the labelled consequence in cases compared to controls. Bars represent the 95% CIs of the point estimates.

## Discussion

In one of the largest exome sequencing studies to date, we identify genes in which disruptive coding variants confer substantial risk for schizophrenia at exome-wide significance. This effort required re-processing a decade of sequence data, harmonization of variant calling and quality control, inclusion of external controls, and integration of PTV, damaging missense, and *de novo* variants. Global, collaborative efforts such as this provide a template for tackling the genetic contributions in other complex diseases.

Genome-wide analyses recapitulated known biological processes and reaffirm that schizophrenia risk genes are involved in the postsynaptic density and broader synaptic function, and enriched in expression in neuronal tissues. Furthermore, the identification of specific genes supports more specific mechanistic hypotheses. The association of PTVs in the NMDA receptor subunit *GRIN2A* to schizophrenia risk provides genetic support for the dysregulation of glutamatergic signaling as a possible mechanism of disease. A natural dose-response curve occurs at this gene in which common regulatory variants modestly influence disease risk and PTV and predicted damaging missense variants increase risk more substantially. Interestingly, the NMDA receptor is composed of two *GRIN2* units *(GRIN2A* and/or *GRIN2B)* along with two constitutive *GRIN1* units, and *GRIN2A* increases dramatically in expression later in childhood and adolescence, mimicking the age of onset of disease for schizophrenia. *De novo* mutations in *GRIN2B* conversely are associated with more severe disorders of neurodevelopment that manifest in childhood, including intellectual disability and autism^41^. Such findings provide a unique opportunity to identify experiments of nature which help to build and support mechanistic hypotheses that may lead to a better understanding of disease biology.

Joint analysis with genetic data from DD/ID and ASD consortia have provided evidence for shared genes between neuropsychiatric and broader neurodevelopmental disorders. Indeed, seven of the 32 FDR < 5% genes are also associated with DD/ID, providing additional confidence in those associations. The shared genes suggest that there is at least some contribution from early brain developmental processes that predisposes to schizophrenia. Despite this sharing, PTVs in 9 of the 10 most confidently associated genes are associated with schizophrenia and not for DD/ID, which may provide avenues for identifying disease-specific processes. Of further interest, we observe allelic series in *GRIN2A, TRIO*, and *CACNA1G* in which PTVs increase schizophrenia risk and *de novo* missense mutations confer strong DD/ID risk. *De novo* missense mutations in these genes clustered in specific domains and are associated with more severe neurodevelopmental, syndromic disorders with cognitive impairment, suggesting an alternate or gain-of-function effect. Analyses estimating relative penetrance for different phenotypes will increase in power as consortium efforts studying specific diseases and biobank efforts continue to grow, all of which would be fruitful in informing what is shared and distinct across disorders.

We show for the first time that common regulatory variants from GWAS and ultra-rare coding variants disrupt an overlapping set of genes, including an allelic series in four genes in which common variants and rare coding variants increase risk to varying degrees. Combined, these results suggest that exome sequencing identifies some common, shared underlying biology that is dysregulated across the allele frequency spectrum, rather than syndromic forms of disease with unrelated biology regulated by common variation. Furthermore, because of this sharing, coding variants can help refine and fine-map common variant associations like at the *STAG1* and *FAM120A* loci. As common and rare variant association studies continue to grow, we can better determine the actual degree of overlap of genes that are regulated by both types of variation. Ultimately, the emerging evidence of an overlap between common and ultra-rare variation gives confidence that the integration of results from sequencing consortia with the GWAS efforts will have significant value for identifying specific genes beyond what any single strategy can achieve on its own.

A decade of genotyping and sequencing studies now establish specific genetic contributions from common variants, copy number variants, and ultra-rare coding variants as conferring risk for schizophrenia. Despite this progress, it is clear that we are still in the early stages of gene discovery^10^. The vast majority of risk alleles, their direction and magnitude of effect, mode of action, and responsible genes are yet to be discovered. These emerging genetic findings will serve in part to direct and motivate mechanistic studies that begin to unravel disease biology. The success of common variant association studies, and now exome sequencing, suggest concrete progress towards understanding the causes of human complex traits and diseases, and provide a clear roadmap towards understanding the genetic architecture of schizophrenia.

#### Box 1

A brief review of literature on exome-wide significant genes.

##### *SETD1A* (SET Domain Containing 1A, Histone Lysine Methyltransferase)

*SETD1A* encodes a catalytic subunit of the COMPASS histone methyltransferase complex that performs mono-, di-, and tri-methylation at Histone H3 Lysine 4. Through epigenetic modification, SETD1A influences transcriptional regulation that downstream affects axonal branching and cortical synaptic dynamics^42^. Paralogs of this gene, including *KMT2A* (Wiedemann-Steiner syndrome), *KMT2B, KMT2C*, and *KMT2D* (Kabuki syndrome), have been associated with severe neurodevelopmental disorders^18,43^.

##### *CUL1* (Cullin 1)

*CUL1* is a core component of a E3 ubiquitin-protein ligase involved in the ubiquitination of proteins broadly involved in signal transduction, gene regulation, and cell cycle progression^44,45^. CUL1, RBX1, and SKP1 form the invariable components of the SCF complex that recruits proteins for degradation^46^. Mouse studies have demonstrated that CUL1 is required for early development, and mutant mice fail to regulate G1 cyclin (cyclin E) during embryogenesis^46^.

##### *XPO7* (Exportin-7)

XPO7 is involved in the trafficking of specific proteins through the nuclear pores to the cytoplasm as part of the Ran-GTP pathway^47^. In a GTP-bound form, exportins bind to nuclear export signal sequences of select proteins, interact with nucleoporins to transit into the cytoplasm, release the cargo protein, and relocate to the nuclear compartment. While exportins are generally involved in nuclear export, XPO7 may be involved in the nuclear import of select substrates as well^47^.

##### *TRIO* (Trio Rho Guanine Nucleotide Exchange Factor)

*TRIO* encodes for a Rho GDP to GTP exchange factor that promote actin cytoskeleton reorganization through the direct activation of the signaling G protein RAC1. TRIO has been demonstrated to regulate neuronal migration, axonogenesis, axon guidance, and synaptogenesis through actin cytoskeleton remodeling. Mutations in *TRIO* are associated with intellectual disability and neurodevelopmental disorders, with mutational hotspots at the seventh spectrin repeat and RAC1-activating GEF1 domain^37,48^. Specific mutations in these domains cause hyper- or hypo-activation of RAC1, resulting in varying severities of developmental delay^37^. TRIO is been shown to directly affect glutamatergic neurotransmission in rodent neurons *in vitro*: specific hypomorphs in the GEF1/Rac1 activating domain reduce synaptic AMPA receptor expression, while hyperfunctional mutations enhance glutamatergic signaling and synaptogenesis^49^.

##### *CACNA1G* (Calcium Voltage-Gated Channel Subunit Alpha-1G subunit)

*CACNA1G*, also known as *Cav3.1*, encodes for the alpha 1 G subunit of a T-type voltage-sensitive calcium channel that mediates calcium influx into excitable cells^50^. T-type channels produce transient and small currents that modulate the firing patterns of neurons and cardiac nodal cells, which has been shown to influence muscle contraction, neurotransmitter release, cell division and growth, cell motility, and sleep stabilization^51,52^. A specific functional mutation in *CACNA1G* has been linked to hereditary cerebellar ataxia in a single family^53^.

##### *SP4* (Sp4 Transcription Factor)

*SP4* encodes a transcription factor that binds to GT and GC promoter elements to activate transcription. SP1 and SP3 transcription factors recognize similar DNA elements, but SP4 has stronger specific abundance in the central nervous system^54^. SP4 knockout mice develop until birth without abnormalities but two-thirds die within four weeks of birth and the remaining third suffer severe developmental delay^54,55^. These mice also experience robust deficits in sensorimotor gating and contextual memory associated with hippocampal vacuolization^56^. Furthermore, SP4 knockout causes dramatically decreased expression of *GRIN1*, the constitutive subunit of the NMDA receptor, which may impact NMDA neurotransmission^57^.

##### *GRIA3* (Glutamate Ionotropic Receptor AMPA Type Subunit 3)

*GRIA3*, or *GluA3*, encodes for a subunit of the tetrametric AMPA-sensitive glutamate receptor, which, as part of the glutamate receptor family, serve the predominant excitatory neurotransmitter receptors in the mammalian brain. AMPA receptors are well-characterized in their involvement mammalian central nervous system function, and moderates fast excitatory synaptic transmission and are involved in long-term potential and synaptic plasticity^58-60^. Individual mutations in two AMPARs, *GRIA2* and *GRIA3*, have been linked with intellectual disability in small patient cohorts^61,62^.

##### *GRIN2A* (Glutamate Ionotropic Receptor NMDA Type Subunit 2A)

*GRIN2A*, also known as *NR2A* or *GluN2A*, encodes for the 2A subunit of the glutamatergic N-methyl-D-aspartate receptors (NMDARs) which, along with AMPARs, are implicated in learning, memory and synaptic plasticity involving long-term potentiation. NMDA receptor dysregulation has been long postulated as a hypothesis for schizophrenia etiology^63^. NMDAR channel blockers (ketamine and phencyclidine) induce schizophrenia-like symptoms and cognitive deficits in healthy individuals^64,65^, and anti-NMDAR encephalitis, an autoimmune disorder originating from a antibody-mediated attack on NMDARs, result in psychosis-like symptoms and memory deficits^66^. Protein-truncating and specific missense mutations in *GRIN2A and GRIN2B* cause neurodevelopmental disorders with prominent speech features and epilepsies^41,67^. A patient cohort of 248 individuals show that missense variants in the transmembrane and linker domains are associated with severe developmental phenotypes, while PTVs and missense variants outside the pore result in mild cognitive defects^68^.

##### *HERC1* (HECT And RLD Domain Containing E3 Ubiquitin Protein Ligase Family Member 1)

*HERC1* is a large E3 ubiquitin protein ligase with HECT and RCC1-like as its characteristic protein domains. *HERC1* is involved in membrane trafficking and cell proliferation through its interactions with clathrin, M2-pyruvate, and TSC2^69–71^. A specific recessive missense mutation in the RCC1-like domain causes severe ataxia and early death through progressive Purkinje cell degeneration^71^.

##### *RB1CC1* (RB1 Inducible Coiled-Coil 1)

*RB1CC1* encodes for a DNA-binding transcriptional factor that coordinates cell proliferation, autophagy, and cell migration^72,73^. It binds to a GC-rich region upstream of the RB1 promoter and forms a complex with p53 to activate RB1 expression. Deletions of *RB1CC1* have increased frequencies in human breast cancers, with its involvement in cancer development primarily attributed through its regulation of RB1^72^.

## Data Availability

We have provided data tables in the main text, Supplementary Methods, and in an exome results browser at https://schema.broadinstitute.org

